# Microstructural deficits in the thalamus of major depressive disorder

**DOI:** 10.1101/2021.10.11.21264842

**Authors:** Yuxuan Zhang, Yingli Zhang, Hui Ai, Nicholas T. Van Dam, Long Qian, Gangqiang Hou, Pengfei Xu

## Abstract

Macroscopic structural abnormalities in the thalamus and thalamic circuits have been shown to contribute to the neuropathology of major depressive disorder (MDD). However, cytoarchitectonic properties underlying these macroscopic abnormalities remain unknown. The purpose of this study was to identify systematic deficits of brain architecture in depression, from structural brain network organization to microstructural properties. A multi-modal neuroimaging approach including diffusion, anatomical and quantitative magnetic resonance imaging (MRI) was used to examine structural-related alternations in 56 MDD patients compared with 35 age- and sex-matched controls. Structural networks were constructed and analyzed using seed-based probabilistic tractography. Morphometric measurements, including cortical thickness and voxel-based morphometry (VBM), were evaluated across the whole brain. A conjunction analysis was then conducted to identify key regions showing common structural alternations across modalities. The microstructural properties, macromolecular tissue volume (MTV) and T1 relaxation times of identified key regions were then calculated. Results showed multiple alterations of structural connectivity within a set of subcortical areas and their connections to cortical regions in MDD patients. These subcortical regions included the putamen, thalamus and caudate, which are predominately involved in the limbic-cortical-striatal-pallidal-thalamic network (LCSPT). Structural connectivity was disrupted within and between large-scale networks, mainly including subcortical networks, default mode networks and salience/ventral attention networks. Consistently, these regions also exhibited widespread volume reductions in MDD patients, specifically the bilateral thalamus, left putamen and right caudate. Importantly, the microstructural properties, T1 relaxation time of left thalamus were increased and negatively correlated with its gray matter volume in MDD patients. The present work to date sheds light on the neuropathological disruptions of LCSPT circuit in MDD, providing the first multi-modal neuroimaging evidence for the macro-micro structural abnormalities of the thalamus in patients with MDD. These findings have implications in understanding the abnormal changes of brain structures across development of MDD.

## Introduction

Major depressive disorder (MDD) is a globally prevalent and burdensome psychiatric illness associated with abnormal changes in several cognitive domains, including attention, memory and emotion processing (1). According to the Global Burden of Disease study, depression is the leading cause of disability worldwide (2). A large body of neuroimaging findings show structural deficits across multiple brain regions and widespread disruptions of connectivity between brain networks in MDD. These findings consistently converge on one predominant circuit involving the limbic, striatal, pallidal, thalamic and cortical structures, which might underlie depressive pathology (3-5). Within the circuit, the thalamus serves as a hub that is functionally responsive to the relay and distribution of afferent signals, while anatomically is interconnected with the prefrontal cortex and amygdala (3). Its reciprocal connections with cortical and subcortical regions enable the thalamus to facilitate exchanges of subcortical information with the cortex (6). However, the specific role of the thalamus in the neuropathology of depression remains unknown.

Neuroimaging findings have also converged on abnormal structural connectivity and volume reductions of the thalamic network in depression. Specifically, diffusion tensor imaging (DTI) studies have shown abnormal structural connectivity and reduced white matter tracts within the thalamo-frontal pathyway in MDD (7, 8). By combining DTI with graph theory, several groups have shown disrupted topological organizations of structural networks in key regions of the thalamic network in MDD (9, 10). Complementary to DTI-based structural connectivity, accumulating evidence has also shown morphometric abnormalities of the thalamic network in MDD. Gray matter volume (GMV) and/or white matter volume (WMV) reduction have been found in the thalamus (11, 12), amygdala (13) and putamen (14). Previous studies have also shown associations between thalamic volume and severity of depressive symptoms (15). Decreased cortical thickness in the prefrontal cortex (PFC) has been observed (16). Together, alterations have been observed across all parts of the limbic-cortical-striatal-pallidal-thalamic circuit (LCSPT). These findings jointly converge upon macroscopic structural abnormalities within the thalamic network (e.g., the LCSPT circuit) that may potentially contribute to the neuropathology of MDD. However, quantitative alternations of cytoarchitectonic-related properties at a micro-structural level of the thalamus, and its connectivity within the LCSPT circuit remain unclear.

By estimating brain macromolecular tissue volume (MTV) and T1 relaxation times, the recently developed quantitative magnetic resonance imaging (qMRI) technique can inform about macromolecular composition and organization (17). Specifically, MTV quantifies the amount of non-proton tissue within a voxel, while quantitative T1 depends on both density of macromolecules and the local microenvironment. Developmental decrease of T1 is thought to be a consequence of microstructural proliferation such as dendrites, oligodendrocytes and myelination (18). The qMRI technique has successfully been applied to identify the brain tissue changes, quantify the microstructural properties of specific regions and monitor disease status in clinical practice (17, 18). For instance, in patients with multiple sclerosis, a reduction in the MTV along the corticospinal tract has been observed, suggesting qMRI measurements could potentially be served as a noninvasive biomarker for disease monitoring in clinical application (17).

Here, we used a multi-modal neuroimaging approach comprising anatomical, diffusion and quantitative MRI to examine integrative structural abnormalities in patients with MDD, from macroscopic structural networks to the cytoarchitectonic-related properties of the LCSPT circuit at the micro-structure level. We first explored abnormalities within- and across-network structural connectivity and the highly connected hub regions, as well as cortical thickness and subcortical volume across the whole brain. Next, we conducted a conjunction analysis to identify key regions that showed common structural alternations within the LCSPT circuit across multi-modal imaging data. Finally, we examined alternations in tissue properties as well as the relationship between tissue properties and macroscopic measurements of identified key regions in MDD.

## Materials and methods

### Participants

In total, 56 MDD patients (aged 36 years, 41 females) and 35 age- and sex-matched healthy controls (HCs; aged 39 years, 22 females) were included in the present study. Demographic and clinical characteristics of all participants were recorded in supplementary Table S1. For further clinical details about the patients, see Supplementary Methods.

### MRI data acquisition

MRI data was collected from all participants (n = 91) using a 3T GE Discovery MR750 scanner (GE Medical Systems) at Shenzhen Kangning Hospital with an 8-channel head coil (see Supplementary Methods).

### MRI data analysis

An overall workflow of MRI data processing is summarized in Figure 1. The workflow is composed by three sub-workflows, separated into diffusion, anatomical and quantitative MRI processing streams.

**Figure 1.**
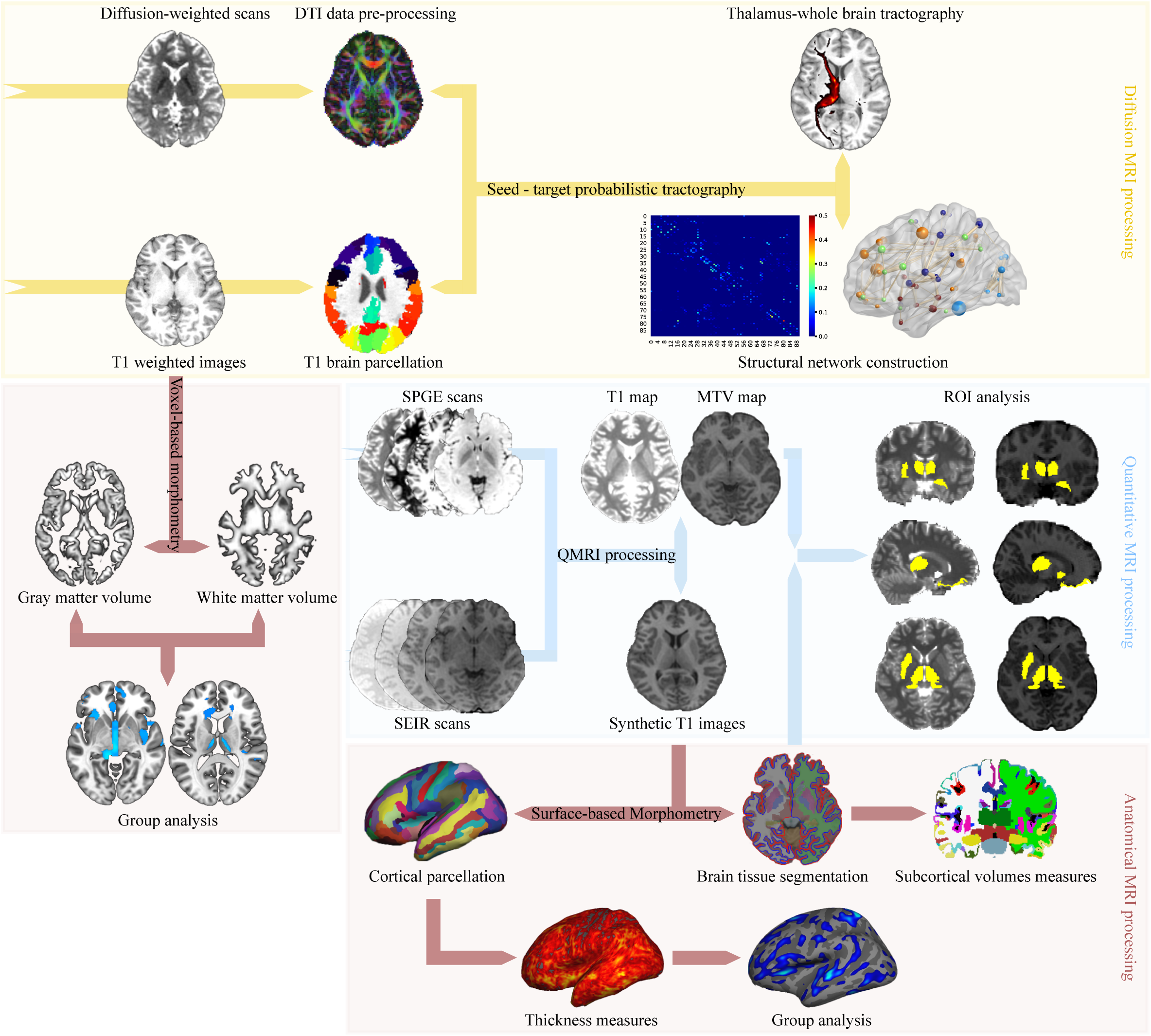
An overview of the MRI data analysis workflow. In diffusion MRI processing stream, the DTI model was constructed from diffusion-weighted images and individual T1-weighted images were parcellated into 90 cortical and subcortical regions according to the AAL atlas. The thalamus-whole brain connectivity and whole-brain structural network were constructed using ‘seed-target’ probabilistic tractography. In quantitative MRI processing stream, individual MTV and T1 maps with synthetic T1 images were generated and used for ROI analysis. In anatomical MRI processing stream, the synthetic T1 images were segmented for qMRI ROI analysis and cortical thickness was calculated. Additionally, individual T1-weighted images were processed for VBM analysis. DTI, diffusion tensor imaging; SPGE, spoiled gradient echo; SEIR, spin echo inversion recovery; MTV, macromolecular tissue volume; VBM, voxel-based morphometry.

#### Diffusion MRI data analysis

The diffusion MRI (dMRI) data were preprocessed followed by structural network construction and analysis to evaluate the whole-brain structural connectivity in MDD patients. An automated anatomical labeling (AAL; 19) atlas was used to parcellate the whole cerebral cortex into 90 regions/nodes (45 regions in each hemisphere, without the cerebellum; supplementary Table S2) in native diffusion space, followed by probabilistic tractography between 90 AAL regions. This resulted in a connectivity matrix with inter-regional connection probability, representing a structural network for each participant. The Network-based statistic (NBS; 20) analysis was performed to determine the difference of inter-regional structural connectivity between MDD and HC groups. Supra-threshold connections at a significant level of *p* < 0.01 after correcting for family-wise error (FWE) were reported. For further details on dMRI data processing, see Supplementary Methods.

#### Anatomical MRI data analysis

A voxel-based morphometry (VBM) analysis using the VBM8 toolbox (http://dbm.neuro.uni-jena.de/vbm8/) combined with surface-based morphometry (SBM) using Freesurfer (http://surfer.nmr.mgh.harvard.edu; 21, 22) was performed to detect morphologic alternations in MDD patients (see Supplementary Methods).

#### Quantitative MRI data analysis

The qMRI data were preprocessed by using the mrQ software package (https://github.com/mezera/mrQ; 17) to produce the evaluation of MTV and quantitative T1 maps. To examine microstructural disruptions underlying the common alterations in structural connectivity and regional morphology, we conducted region of interest (ROI) analyses on qMRI measurements of the regions identified in conjunction analysis (see Supplementary Methods).

We conducted Pearson correlation analyses to examine the relationships between microstructural properties (MTV and T1) and two other measurements, the GMV and fractional anisotropy (FA) within key regions identified by conjunction analysis. For details, see Supplementary Methods.

### Statistical analysis

Statistical analyses were performed using IBM SPSS (version 20). The Chi-square test and *t*-test were conducted to compare potential between-group differences in gender, age and education, with a statistical significance of *p* < 0.05. For the volumes of subcortical regions, a general linear model (GLM) was conducted controlling for age, sex and education years, with a statistical significance of *p* < 0.05 after False Discovery Rate (FDR) correction. For ROI analysis of qMRI data, a two-sample *t*-test was conducted with participants’ age and gender as covariates to test group difference in MTV and T1 within each ROI. Significance was set at *p* < 0.05, FDR-corrected.

## Results

### Demographic and clinical characteristics

There were no significant differences in age nor gender between MDD patients and HCs (age: *t* (89) = -0.910, *p* = 0.365; gender: χ^2^ (1) = 1.085, *p* = 0.353). There was a significant group difference in years of education (*t* (89) = -3.228, *p* = 0.002). To test whether our results of MRI data analyses were affected by the demographic data, we separately repeated the MRI data analyses with and without the demographic data as covariates, and we compared the different results to evaluate the potential influences of demographic variables.

### Alternations of structural connectivity in MDD patients

The NBS analysis showed two networks with disrupted structural connectivity in patients with MDD (Figure 2). Specifically, a network comprised of 215 edges that connected to 76 nodes, showing significantly increased structural connectivity in MDD (Figure 2A, MDD > HC). Another network consisted of 44 edges linking to 38 nodes, showing significant decreased structural connectivity in MDD (Figure 2B, MDD < HC). For illustrating the connectivity patterns among large-scale networks in patients with MDD, the identified increased and decreased structural connectivity were respectively categorized into a well-known seven-network parcellation of the human cerebral cortex with 100 parcels (Figure 2C and D; 23). Given the importance of subcortical regions in our study, we included the subcortical regions as an additional network division. The AAL regions were localized into the seven networks based on their Montreal Neurological Institute (MNI) coordinates. We found that the most increased and decreased structural connectivity were those within the subcortical regions and those between the subcortical regions and cortical networks, including the default mode network (DMN) and salience/ventral attention network (SN/VAN). The hub regions that were involved in more than ten inter-regional connections were summarized as bar plot in Figure 2E. For details on alternations of structural connectivity in MDD patients, see Supplementary Results.

**Figure 2.**
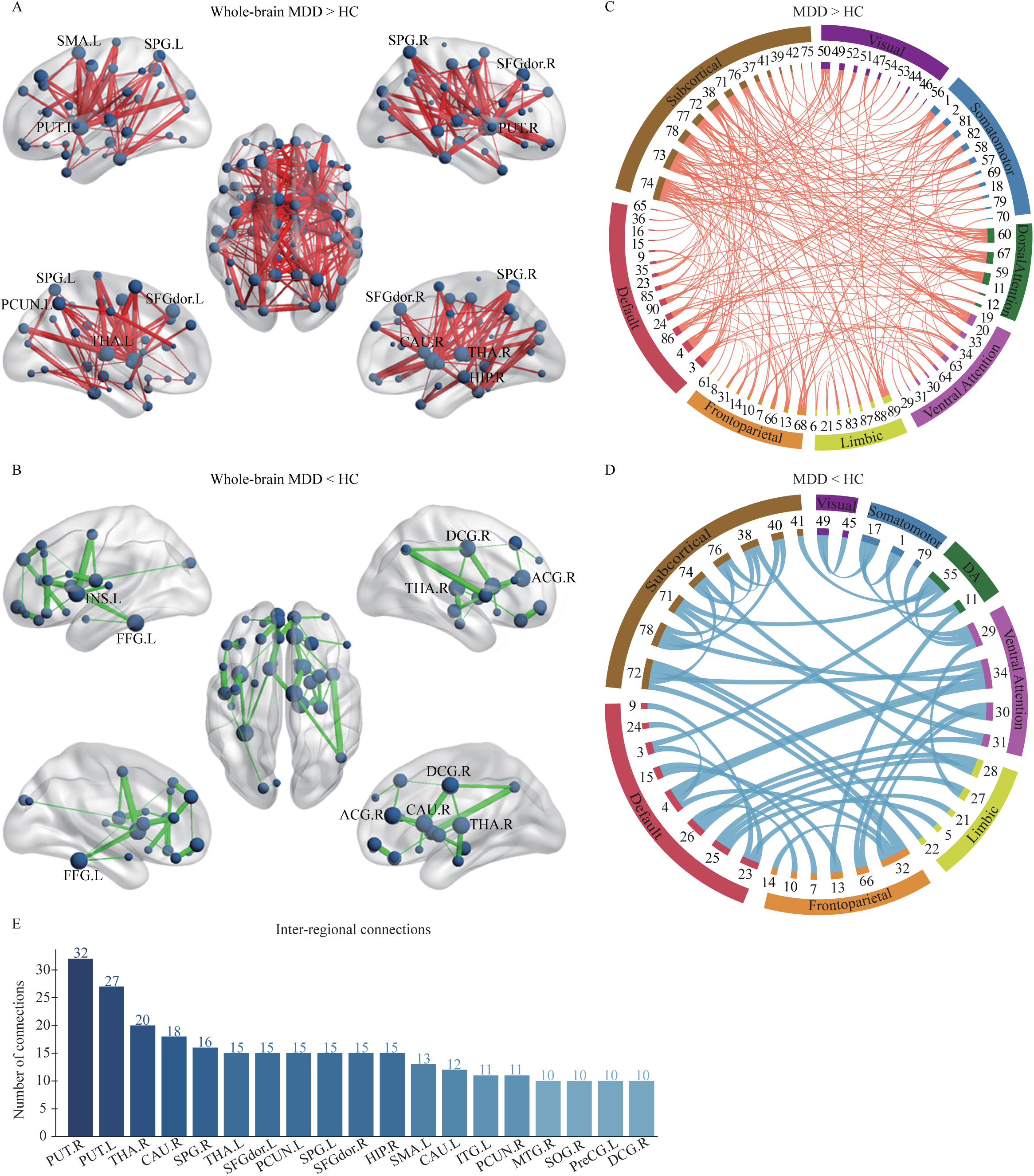
Altered structural connectivity in MDD patients. (A) Increased and (B) decreased connectivity identified by the NBS analysis in MDD patients relative to HCs. All nodes are shown in dark blue with node sizes indicating number of edges the given nodes involved. The nodes with labels are the hub regions. The edges representing increased and decreased connections are shown in red and green, with edge widths representing *t* values between nodes. The circle plots depicting the increased (C) and decreased (D) connectivity patterns within and between 7 networks and subcortical networks. AAL regions are represented by the inner segments of circle plots and abbreviated and labelled as their index in AAL atlas. All the regions are circularly arranged according to their number of connections and the angular size of each inner segment is proportional to the total connections that given region involved. Networks are labelled on the outer segments. Regions within the same networks are shown in the same color. The *t* values of connections are represented by ribbons between the inner segments. The ribbons for increased connections are shown in red and for decreased connections in blue. (E) The hub regions that involved in more than 10 inter-regional connections are sorted in a descending order. DA, Dorsal Attention. For abbreviation and index of AAL regions, see supplementary Table S2.

### Alternations of GMV/WMV in MDD patients

Significant group differences were observed in both GMV and WMV (Figure 3 and supplementary Table S3). The reduced GMV/WMV of these regions in MDD was not substantially altered after regressing out covariates (see Supplementary Results). There was no cluster where GMV or WMV was significantly larger in MDD patients than those in HCs.

**Figure 3.**
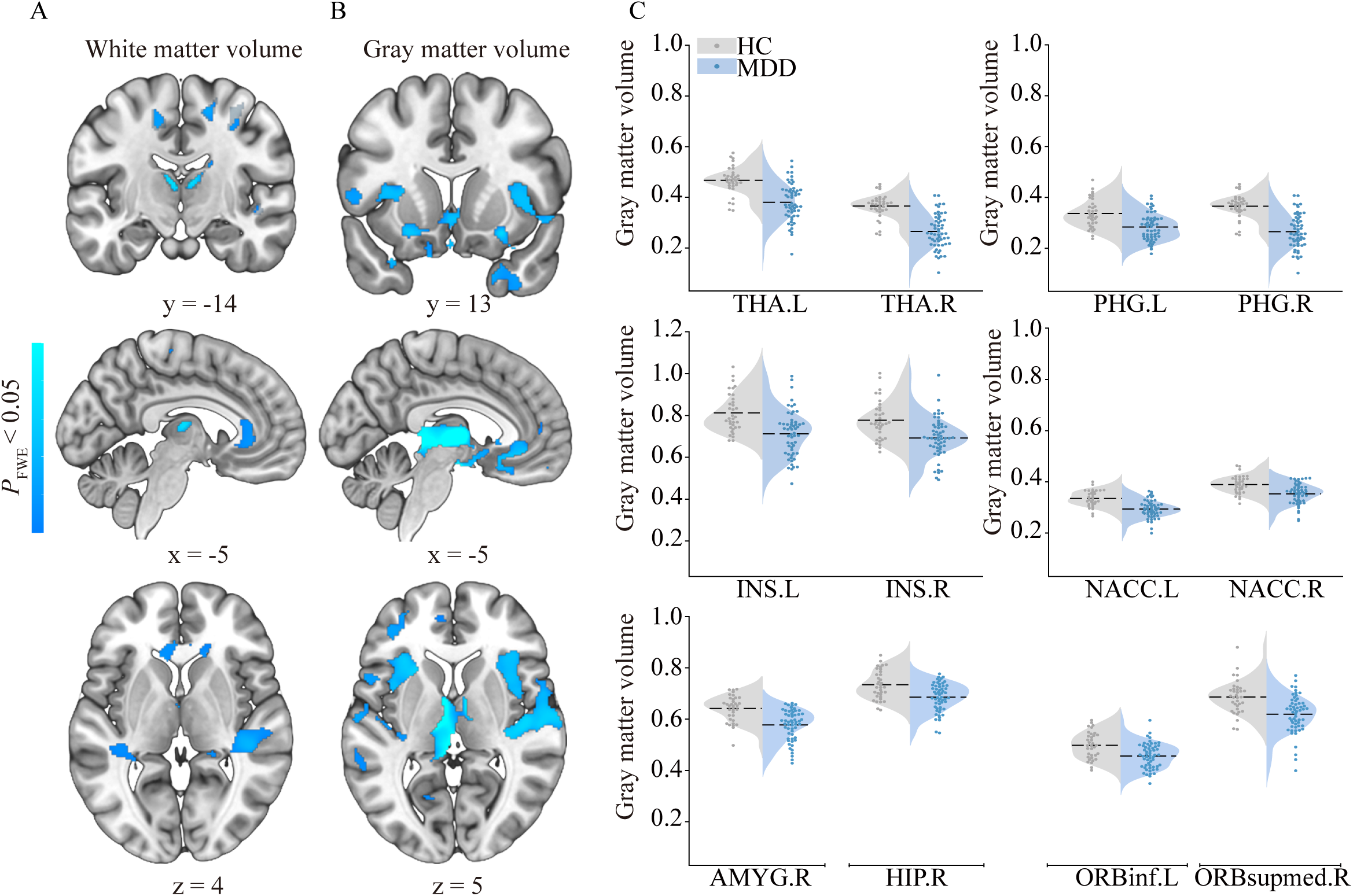
Between-group differences in the WMV and GMV. (A) WMV and (B) GMV reductions in MDD patients compared with HCs. The color bar corresponds to FWE-corrected *p* < 0.05 and lower. Representative coronal, sagittal and axial slices of the supra-threshold clusters were overlaid on a MNI152 template. (C) Violin plots depicting between-group differences of the GMV values within 12 key regions, where the width indicates subject density, dashed line indicates average gray matter volume and scatters indicate subject distribution for each group. Since the raw statistic map was transformed into TFCE values after TFCE method, the GMV values of these regions were derived from the group-comparison statistic map with threshold of *p* < 0.001 (uncorrected) at the voxel level. For abbreviation of AAL regions, see supplementary Table S2. NACC, nucleus accumbens; GMV, gray matter volume; WMV, white matter volume.

### Results of SBM analysis

We found 14 clusters where cortical thickness was significantly lower in MDD patients relative to HCs (Figure 4 and supplementary Table S4). No significant increases in cortical thickness were detected in MDD group. None of the subcortical volume showed significant group differences after FDR correction.

**Figure 4.**
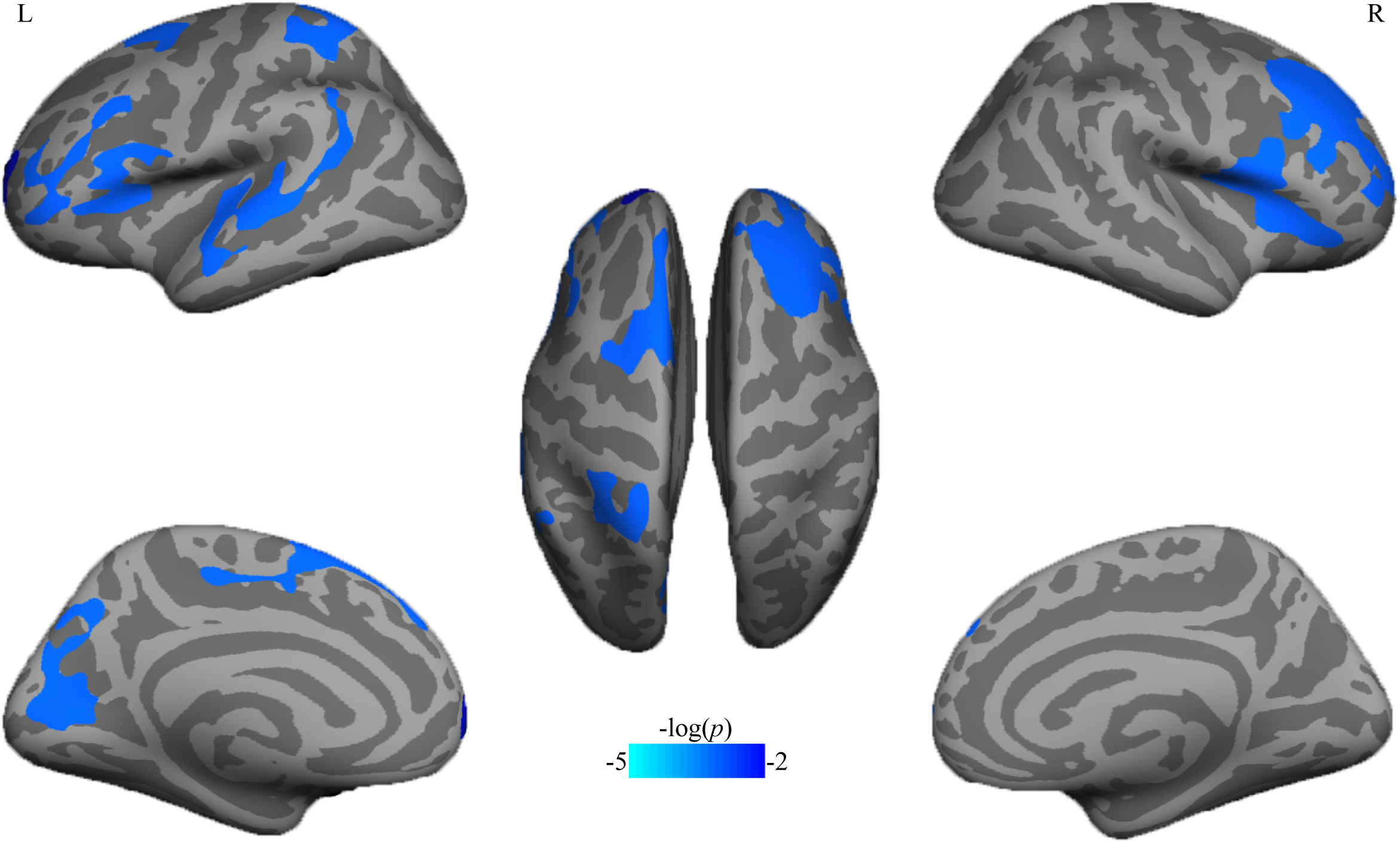
Cortical thickness changes between MDD patients and HCs. Significant differences in cortex thickness were mapped onto the bilateral inflated cortical surfaces at the lateral and medial views. Dark gray indicates gyri; light gray indicates sulci. The color bar represents the *p* value. L, left hemisphere; R, right hemisphere.

### Results of qMRI analysis

We found averaged T1 values were significantly higher in MDD patients than HCs in the left thalamus (THA.L; *t* (89) = 2.687, FDR-corrected *p* = 0.045; Figure 5B). This result was not significantly correlated with participants’ education (*r* = -0.193, *p* = 0.067). No significant between-group difference in qMRI measures was observed in other ROIs after FDR correction.

**Figure 5.**
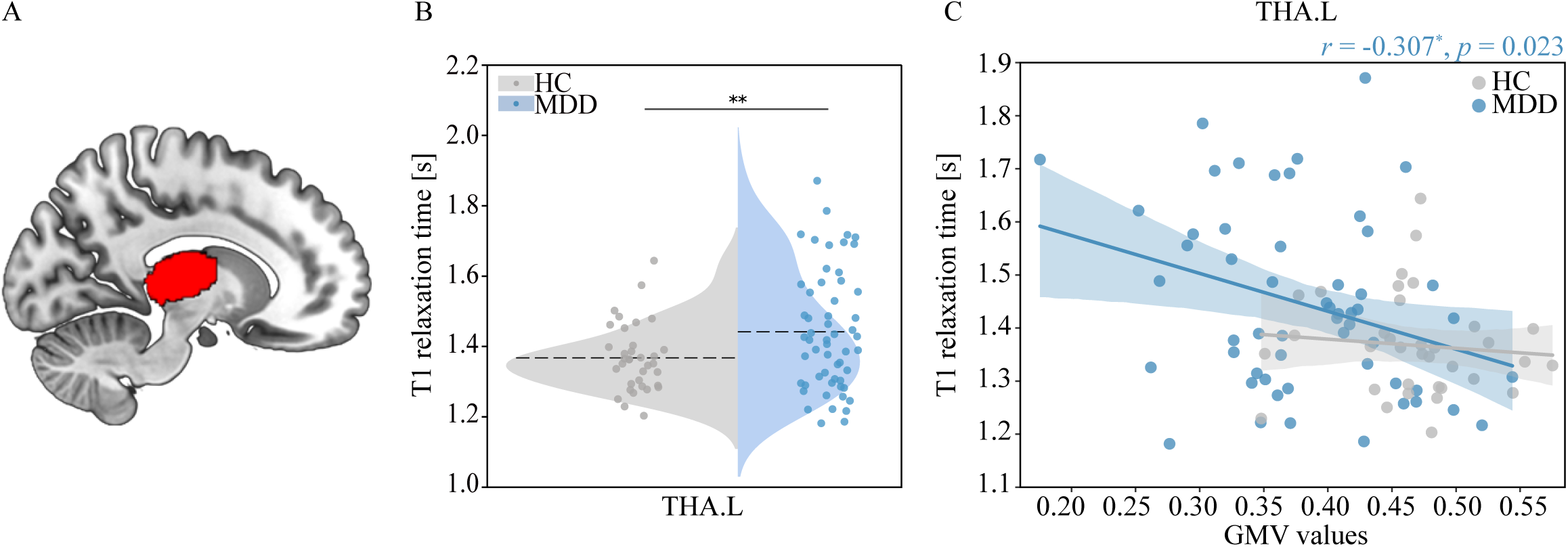
Group differences in T1 values and the correlation with GMV. (A) The volumetric mask of the left thalamus was mapped on a MNI152 template for visualization. (B) The Violin plot shows between*-*group differences in T1 relaxation times in the left thalamus. (C) Correlation between GMV and T1 values in the left thalamus in MDD group. The blue dots represent patients with MDD while the gray dots represent HCs. The correlation coefficient (*r*) and *p* value in MDD group was shown in blue and labelled on the upper right of the plot. Note that the left thalamus was segmented based on Destrieux Atlas in individual space (see *Anatomical MRI data analysis*). MDD, major depression disorder; HC, healthy controls. For abbreviation of AAL regions, see supplementary Table S2.

The correlation analysis of qMRI measurements showed that T1 values were negatively correlated with GMV values in the THA.L (*r* = -0.307, FWE-corrected *p* = 0.023; Figure 5C) specifically in MDD but not in HCs. There was no significant correlation observed between T1 and FA values in the THA.L in MDD group (*r* = -0.109, FWE-corrected *p* = 0.423).

## Discussion

To our knowledge, this is the first study combining multi-modal neuroimaging techniques to characterize microstructural abnormalities of the neural pathway that underlies the neuropathology of MDD. Three main findings are revealed in the present study. First, disrupted structural connectivity within and between large-scale networks are closely associated with subcortical regions involved in neural circuits of MDD. Second, hub regions that are highly interconnected in structural networks consistently show morphological abnormalities. Third, the thalamus that plays a key role within the LCSPT circuit exhibits both common macrostructural alternations and cytoarchitectonic-related abnormalities in MDD. These findings elucidate the primary role of the thalamus in macro-to-micro structural abnormalities of depression.

### Disrupted structural connectivity within and between large-scale brain networks

We found deficits of structural connectivity in patients with MDD were characterized by abnormal increased and decreased connectivity within subcortical regions and between the subcortical regions and regions within the DMN and SN/VAN. Importantly, hub regions that interconnected within and between networks were primarily localized within the subcortical regions, specifically the bilateral thalamus, bilateral putamen and right caudate, and also in the bilateral dorsolateral superior frontal gyrus within the DMN, even after controlling for age, gender and years of education. Consistent with previous findings (7, 9, 10), these findings point to the major roles of hub regions especially those from the subcortex in disrupted brain connectivity. More importantly, these structures are highly connected, responsible for the normal regulation of mood, and involved in a variety of neuroanatomic circuits that may underlie the pathophysiology of MDD (3, 4). For example, disrupted white matter (WM) connectivity within the frontal-subcortical networks (7), as well as decreased WM tracts in the frontothalamic loops have been found in depressed patients (8). Closely related to these hub regions is also the LCSPT circuit formed by connections between the PFC, amygdala, striatum, thalamus and pallidum, which is known for its central involvement in mood disorder like depression (3-5).

Among this circuit, the thalamus is critical for cognitive and emotional processes through its higher order relay. Among the higher order thalamic relay nuclei, the mediodorsal thalamic nucleus (MD) has reciprocal connections with the PFC, relaying substantial subcortical projections from the amygdala and parahippocampal cortex to the orbital and medial prefrontal cortex (OMPFC; 3, 5). Given its broad connections with cortical and subcortical structures, growing evidence suggests mapping thalamic connectivity would allow the characterization of distributed circuits abnormalities in neuropsychiatric disorders (24). In this context, our findings showed the PFC was predominantly involved in increased connections with the thalamus in MDD patients, including the bilateral dorsolateral superior frontal gyrus and inferior orbitofrontal gyrus, as well as key components of the LCSPT circuit including the bilateral putamen, caudate and pallidum, as indicated by the thalamus-whole brain tractography. Our findings provide new detail regarding disruption of structural connectivity within the LCSPT circuit in MDD, suggesting thalamic dysconnectivity may be a neural marker for the LCSPT circuit abnormalities in MDD.

At the network level, a lot of evidence has shown abnormalities in functional connectivity within the DMN and SN/VAN in depression (25, 26). By synthesizing these findings, a triple network model has been proposed that particularly emphasizes the roles of the DMN and SN/VAN as well as their key nodes in abnormal patterns of functional connectivity in the depressed brain (27). However, the patterns of anatomical connectivity within and between these large-scale brain networks in MDD still remain unclear. Our findings revealed the DMN and SN/VAN also exhibited disrupted structural connectivity in patients with MDD, of which some key nodes including the precuneus of DMN and the insula of SN/VAN were involved in most of abnormal connections with nodes of other networks especially those from subcortical e.g., the putamen, thalamus, caudate and hippocampus. We suggest these key nodes may represent structural hubs particularly sensitive to network-level disruptions in MDD due to their involvement in altered structural connectivity. Notably, the structural hubs we identified are consistent with functional hubs based on previous evidence of functional connectivity in depression (28), suggesting key nodes of those networks may have tightly link with disruptions of both structural and functional connectivity in depression. Taken together, our findings complement previous work from a structural perspective by using structural connectivity to characterize large-scale brain networks in depression.

### Abnormal morphometry alternations of the depressed brain

Morphometric measurements revealed that abnormal morphologic alterations in the depressed brain were primarily localized in the subcortical regions and tightly associated with those hub regions in structural networks. Volume losses in the bilateral thalamus, right amygdala, right hippocampus, bilateral parahippocampal gyrus, left putamen and bilateral dorsolateral superior frontal gyrus, as well as cortical thickness decreases in the orbitofrontal cortex (OFC) and prefrontal regions were observed in patients with MDD. Numerous studies have shown widespread GMV/WMV reduction in patients with MDD, especially the hippocampus, which is one of the most predominant findings that consistently shows volume reduction (13, 29), suggesting hippocampal abnormalities may be a contributor to the pathogenesis of depression. Previous studies have also shown volume reductions as well as cortical thinning within the PFC and OFC in MDD patients (30, 31). Consistent with these findings, we found the reduced GMV of right hippocampus, reduced PFC and OFC volume and decreased OFC thickness in both hemisphere in patients with MDD, suggesting crucial roles of these brain regions in neuropathology of depression.

Volume losses in subcortical structures within the LCSPT circuit have also been associated with the pathology of MDD (32), and has been proposed to be one of the common markers of MDD (14). For example, volume reduction of the thalamus was thought to help account for its maladaptive bottom-up influence in biased processing of negative stimuli of MDD (15), and therefore is considered to be a potential marker of MDD. However, evidence for volume reductions of those structures is still lacking. Our findings instead showed both GMV and WMV of the bilateral thalamus and GMV of the left putamen were reduced in MDD patients, which to date confirm the morphologic abnormalities of the thalamus and putamen in MDD.

Widespread volume reductions in other subcortical structures of the LCSPT circuit involving the amygdala and caudate have been observed in MDD patients, which support the roles of subcortical structures in the pathophysiology of MDD. However, reduced GMV of the amygdala we observed may be contradictory to most previous studies in which MDD patients showed increased amygdala volumes as compared to HCs (33). One may reason might be attributable to heterogeneity of MDD patients, such as differences in medication status and the number of episodes. Crucially, our findings showed both morphologic abnormalities and disrupted structural connectivity were within the bilateral thalamus, right hippocampus and left putamen, which indicate that the subcortical structures of the LCSPT circuit may largely characterize the overall structural abnormalities in MDD patients.

### Abnormal microstructural alternations of key regions

Our study has revealed for the first time that cytoarchitectonic-related properties of the thalamus was altered in the depressive brain, while the thalamus was also involved in macroscopic structural abnormalities including disrupted structural connectivity and volume reductions in MDD patients. As aforementioned, the structural abnormalities of thalamus such as volume loss are thought to be associated with its dysfunction in MDD. Understanding how the thalamus was involved in macro-micro anatomical changes may have implication for understanding the neuropathology of MDD.

Given T1 decrease has been associated with developmental change of brain tissue (18), larger T1 of the left thalamus in our study reflect that the microstructural properties of left thalamus may be involved in abnormal changes during development in MDD patients. However, decreased T1 could be driven by multiple factors associated with tissue development, such as microstructural proliferation and myelination (18, 34). The myelin volume which appears to increase within the cortex across development is a likely source of T1 changes. Since it linearly contributes to MTV (18), tissue changes that arise from myelination could be indirectly reflected by MTV and T1. However, we did not find any group difference in MTV of the left thalamus between MDD patients and HCs. It may suggest there may be other contributions besides the myelination that lead to microstructural alternations of this region. Future studies are necessary to examine how MTV or T1 related to abnormal changes of brain tissue in MDD.

Although the conventional measurements like GMV derived from VBM are affected by many biological factors, there are potential benefits of combining GMV and qMRI metrics to clarify the contributions of different biological properties (17). Here, we found a negative correlation between T1 and GMV values of the left thalamus in MDD patients, which provides additional information for inferring microstructural properties. Given an inverse relationship between MTV and T1 (17), the negative relationship between T1 and GMV may indicate that the macromolecules of left thalamus such as cell membranes and proteins were partly decreased in MDD patients. Previous findings also have shown development could create new tissue that displaces water, resulting in higher MTV and FA (34). However, we did not observe any correlation between T1 and FA of the left thalamus, which may result from many biological factors such as axon diameter, axion density and myelin-sheath thickness (35). Collectively, these findings show the microstructural properties of the thalamus are closely associated with its macroscopically structural abnormalities in MDD patients, and provide new insights to the relationship between microstructural properties and structural abnormalities in MDD.

Several limitations of the present study should be noted. One main limitation was that patients were medicated. As we did not evaluate the effect of medication use on current findings, we cannot make strong inferences about how macroscopic and microstructural alternations of the thalamus might be affected by medication use. Thus, future studies are necessary to replicate the findings across first episode, unmedicated patients and to establish whether medication use has potential effects on structural alternations of the thalamus. In addition, the MDD group and HCs were not matched for education year, although our results were largely unchanged after controlling for participants’ demographic variables. Another limitation may come from the AAL atlas we used in brain parcellation. Recent studies have suggested that the node definition by different parcellations would produce different connectivity patterns of brain networks (36). Future studies can validate consistency of structural connectivity across different parcellation schemes. Finally, the sample size was not highly consistent between groups, which may result in potential inaccuracy in statistical analysis.

## Contributors

Pengfei Xu and Gangqiang Hou designed research. Gangqiang Hou and Yingli Zhang collected data. Yuxuan Zhang analyzed data. Yuxuan Zhang and Pengfei Xu led the writing of this article. All authors approved the final version for submission.

## Data sharing

The datasets reported in the current study and data analysis scripts are available upon request from corresponding author.

## Acknowledgement

This work was supported by the National Natural Science Foundation of China (31920103009 and 31871137), the Major Project of National Social Science Foundation (20&ZD153), Young Elite Scientists Sponsorship Program by China Association for Science and Technology (YESS20180158), Guangdong International Scientific Collaboration Project (2019A050510048), Guangdong Key Basic Research Grant (2018B030332001), Shenzhen-Hong Kong Institute of Brain Science-Shenzhen Fundamental Research Institutions (2019SHIBS0003), Shenzhen Science and Technology Research Funding Program (JCYJ20180507183500566), Shenzhen Fund for Guangdong Provincial High-level Clinical Key Specialties (No. SZGSP013), and Shenzhen Key Medical Discipline Construction Fund (No.SZXK041).

## Figure Legends

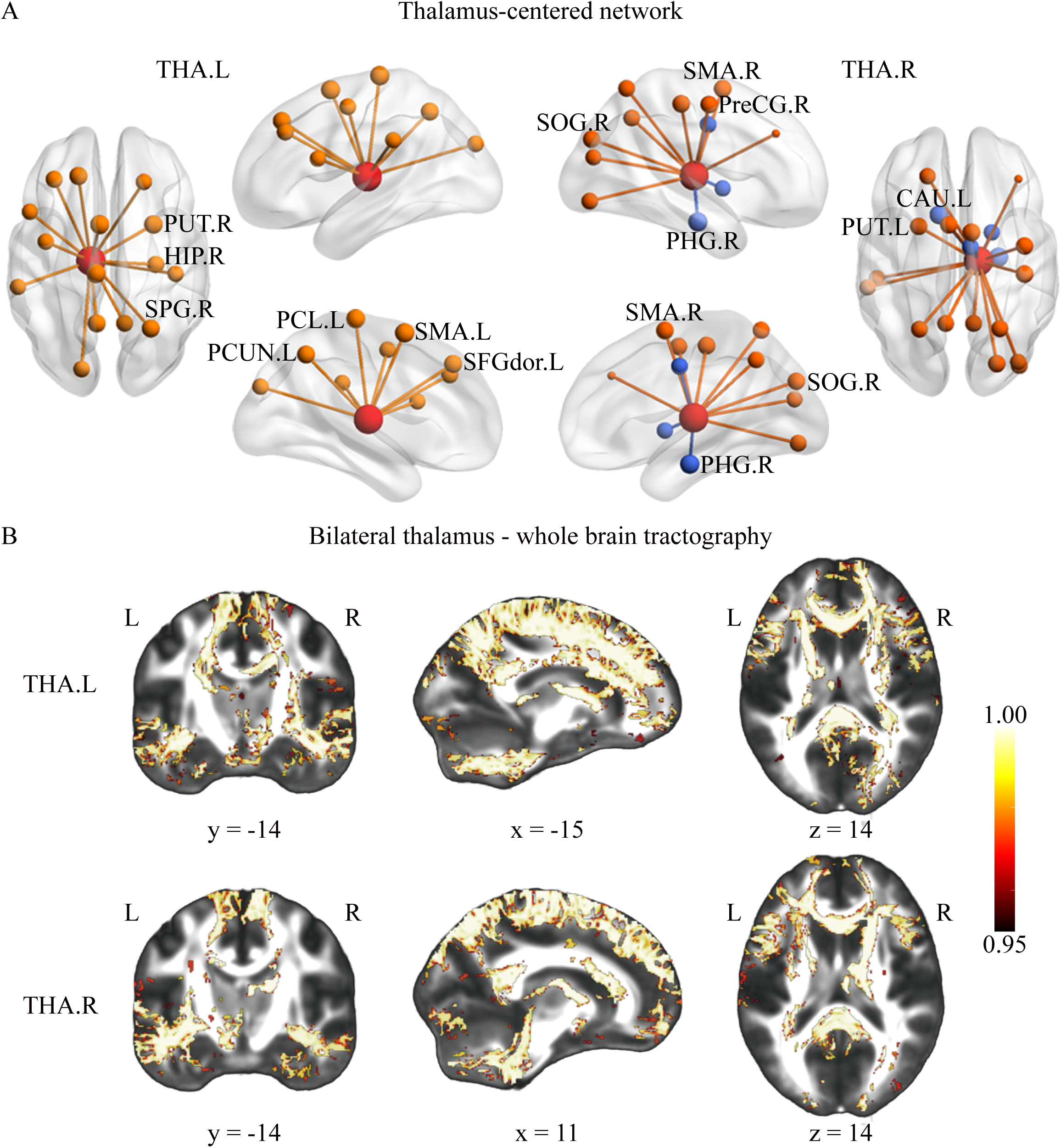

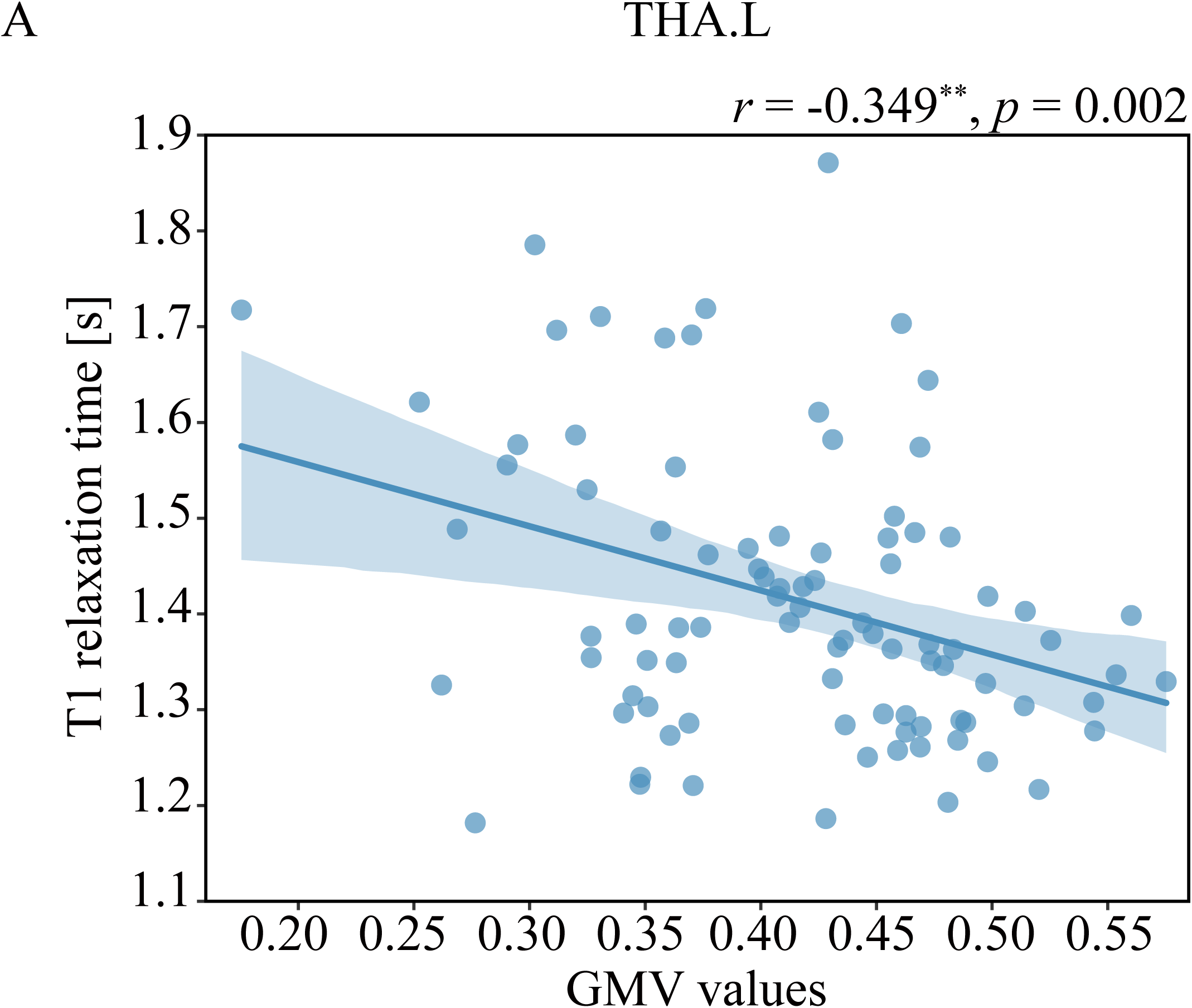

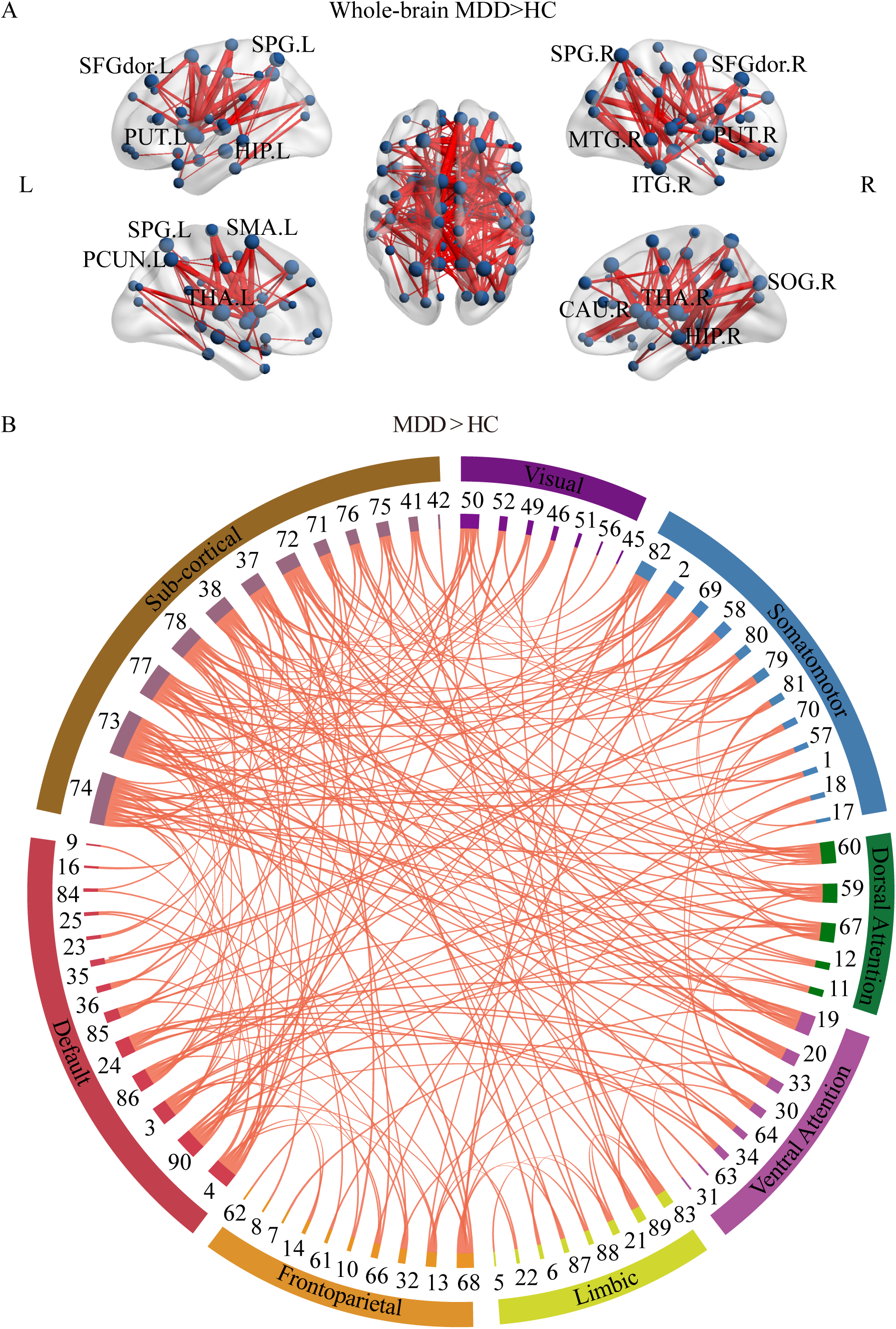

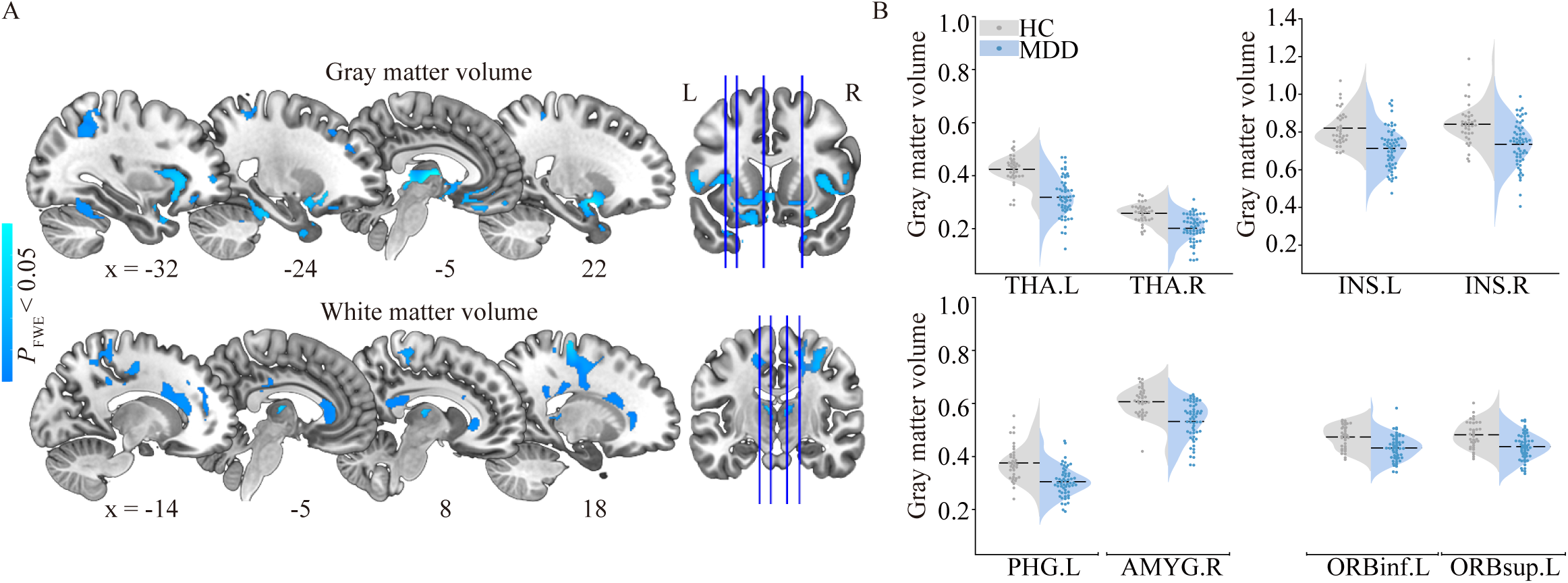

