## Supplementary Materials for "Microstructural deficits in the thalamus of major depressive disorder"

**Supplementary material**

### Supplementary Methods

#### Participants

MDD patients were recruited from Shenzhen Kangning Hospital, diagnosed based on the Structured Clinical Interview for Diagnostic and Statistical Manual of Mental Disorders, Fifth Edition (DSM-V) criteria, and met the following criteria: 1) age range of 18 to 65 years; 2) for male/female who is capable of fertility, s/he agreed to use effective contraceptive methods during the study period and within one month after the end of the study to ensure the effective contraception for himself/herself or sexual partner. The exclusion criteria were as follows: 1) participants met the DSM-V diagnostic criteria for another psychiatric illness, including schizophrenia, bipolar disorder, anxiety disorders, obsessive-compulsive disorder, physical symptoms, organic mental disorders or depression caused by hypothyroidism, substance abuse or dependence; 2) participants who have received electroconvulsive therapy (ECT) within three months before screening; 3) participants with any other unsuited conditions considered by investigators that could not participant the study. The severity of depressive symptoms was assessed using the Hamilton Depression Rating Scale (HAMD) and the severity of anxiety was assessed using the Hamilton Anxiety Rating Scale (HAMA). All HCs were cognitively normal, free of neurological disease, and had no cognitive complaints. All participants and/or the guardians of the MDD patients had signed the informed consent before participating in the study. The study was approved by local ethics committee. For MRI data quality checks, data of one HC were excluded from diffusion MRI (dMRI) data analysis due to the pool image quality, resulting in 56 MDD patients and 34 HCs included in the dMRI data analysis.

#### MRI data acquisition

High-resolution structural images were acquired with a three-dimensional (3D) T1-weighted (T1w) fast spoiled gradient-echo (FSPGR) sequence with TE = 2.9 ms, TR = 6.7 ms, FA = 12°, field of view (FoV) = 256 × 256 mm, 192 slices, and voxel size = 1 mm^3^. The dMRI data was collected using a single-shot spin-echo echo-planar imaging (EPI) sequence (TR = 8724 ms, TE = 81.4 ms, FA = 90°, voxel size = 2 × 2 × 2 mm^3^). The diffusion weighting was isotropically distributed along 64 directions (b = 1000 s/mm^2^). Ten non-diffusion weighted (b = 0) images were acquired at beginning of the dMRI session. The quantitative MRI (qMRI) data was collected using the protocols described in a previous study (1). In brief, the quantitative MTV and T1 relaxation times were measured from four spoiled gradient echo (SPGE) images with flip angles of 4°, 10°, 20°, 30° (TR = 14 ms, TE = 2 ms) at a 1 × 1 mm^2^ in-plane resolution with a slice thickness of 2 mm. We collected four additional spin echo inversion recovery (SEIR) scans with an EPI read-out, a slab inversion pulse, and spectral spatial fat suppression to remove field inhomogeneities. The SEIRs were acquired with a TR of 3 s, echo time set to minimum full, and 2x acceleration. The inversion times were 50, 400, 1200, and 2400 ms with a 2 × 2 mm^2^ in-plane resolution and a slice thickness of 4 mm.

#### Diffusion MRI data analysis

##### Data preprocessing

The dMRI data were preprocessed using the FMRIB’s Diffusion Toolbox (FDT) from FSL package (version 5.0.9; <https://fsl.fmrib.ox.ac.uk/fsl>; 2, 3). Briefly, diffusion-weighted images were first corrected for eddy-current-induced distortions and head movements by using affine transformation of each diffusion-weighted image to non-diffusion-weighted (b = 0) image. The diffusion tensor models were then fitted at each voxel of the brain on distortion-corrected images to yield three eigenvectors and three eigenvalues.

##### Structural network construction

To construct structural brain networks, an automated anatomical labeling (AAL) atlas was used to parcellate the whole cerebral cortex into 90 regions/nodes (45 regions in each hemisphere, without the cerebellum; supplementary Table S2). The mask for each brain region was extracted and transformed into each participant’s native space. In brief, ac-pc aligned T1w image for each participant was first normalized to the ICBM 152 template in Montreal Neurological Institute (MNI) space by applying nonlinear transformation between T1w structural space and standard space. Second, the T1w images in individual participants’ space were then coregistered to their non-diffusion-weighted (b = 0) images by using linear transformation between diffusion space and the T1w structural space. Here, relevant transformation matrices and their inverses were derived and concatenated to produce transformation matrices between diffusion and standard space. The concatenated transformation matrices then used to warp AAL labels from MNI space to native diffusion space. This allowed us to perform inter-subject comparison in diffusion space and produce an individual-specific connectivity matrix. To this end, 90 parcellated regions for each participant in native diffusion space were produced, representing 90 nodes in structural network, and used for structural network analysis.

##### Network analysis

To reconstruct the structural network of white matter tracts between 90 AAL regions, probabilistic tractography was performed on the preprocessed dMRI dataset using PANDA pipeline (<http://www.nitrc.org/projects/panda>; 4) in MATLAB2018b. We used a within-voxel probabilistic diffusion model to build up distributions on diffusion parameters at each voxel of the dataset (5). These probability distributions estimated the multi-fiber tract orientation as well as its uncertainty in each voxel and guided multiple fiber samples starting from a seed voxel to a specified target region. Probabilistic tractography was then applied by drawing 5000 individual streamlines (maximum step: 2000, step length: 0.5mm, curvature threshold: 0.2) in each voxel in the native diffusion space. Structural connectivity between two regions was measured by calculating connectivity probability from each seed region to each of the other target regions. In the present study, each AAL region was selected as the seed and the remaining 89 regions as targets. The tracts were terminated as soon as leaving the brain or reaching a particular target region. This resulted in a 90 × 90 connectivity matrix with inter-regional connection probability, representing a structural network for each participant. For this matrix, the probability from the i-th region to the j-th region is not necessarily equivalent to the one from j to i. However, these two probabilities are highly correlated across the cerebral cortex. Thus, unidirectional symmetric matrix for each participant was acquired by averaging these two probabilities. In the native diffusion space, the resulted structural network composed of nodes, representing a pair of regions, and edges, representing structural connectivity among the nodes, i.e., the region (i) and another region (j) were connected through an edge (e_ij_ = [i, j]), in case of at least one fiber existed between them (6, 7). Consequently, given the definition of nodes and edges for the structural connectivity network (matrix), the number of fibers connecting a pair of regions was calculated as an edge for each participant.

##### Network-based statistics analysis

The NBS approach was applied for group comparisons by using NBS Connectome (<http://www.nitrc.org/projects/nbs/>). The NBS is a non-parametric statistical method to deal with the multiple comparisons problems by controlling the FWE rate in a weak sense while performing mass univariate hypothesis testing on all connections. With the NBS, the null hypothesis was evaluated at a level of interconnected subnetworks rather than individual connections (8). Briefly, a two-sample *t*-test at each edge was conducted independently to test for significant differences in the value of connectivity between two groups. A primary threshold (*t* ≥ 2.5, which corresponds to uncorrected *p* ≤ 0.01) was then applied to define a set of supra-threshold connections. This step identified all the possible connected components in the matrix at this uncorrected level, and followed by computing the size of connected components which link supra-threshold connections with set of nodes in the network. To estimate the statistical significance of size for each component, the null distribution of maximal connected component size was empirically derived using non-parametric permutation testing with 5000 times. Supra-threshold connections that were significant at level of FWE-corrected *p* < 0.01 were reported. The BrainNet viewer (<http://www.nitrc.org/projects/bnv/>; 9) was used to visualize and display the significant connections.

##### Structural connectivity analysis of the thalamus with whole brain.

A seed-based probabilistic tractography was conducted to evaluate white matter structural connectivity of the thalamus with the whole brain in patients with MDD. The steps followed similar procedures with the seed-target probabilistic tractography as described above (see **Network analysis**) but only each unilateral thalamus was defined as a seed and voxels of the rest of the whole brain were set as targets. The unilateral thalamus seed mask adapted for the analysis was derived from the individual T1 parcellation based on AAL atlas. A two-sample *t*-test was conducted to compare the thalamus-centered structural connectivity between MDD patients and HCs. Multiple comparison correction was performed using threshold-free cluster enhancement (TFCE; 10), a non-parametric method to enhance detectability of neuroimaging signal on cluster-based thresholding by integrating information of cluster extend and height into voxel-wise statistical inference. Significant level was set at cluster-wise *p* < 0.05, FWE-corrected.

#### Anatomical MRI data analysis

##### Voxel-based morphometry analysis

To detect macroscopic abnormalities of brain volumes in MDD, VBM analysis was performed using the VBM8 toolbox (<http://dbm.neuro.uni-jena.de/vbm8/>) implemented in SPM 12 (Statistical Paramertic Mapping, <http://www.fil.ion.ucl.ac.uk/spm/software/spm12>) software running on MATLAB R2018 for Linux. All ac-pc aligned T1-weighted images were segmented into gray matter (GM), white matter (WM) and cerebrospinal fluid (CSF) in native space. The segmented images were then normalized to MNI space with a dimension of 121 × 145 × 121 and a spatial resolution of 1.5 × 1.5 × 1.5 mm using high-dimensional DARTEL normalization. The normalized segmented images for GM and WM volume were subsequently modulated by Jacobian determinants derived from the spatial normalization, which allows to estimate regional cerebral volume while controlling for overall brain size. After preprocessing, sample homogeneity was checked to identify outliers which were two standard deviations outside of the mean. Finally, these normalized, segmented, modulated volumes were spatial smoothed with a 6-mm full-width half maximum (FWHM) Gaussian kernel. To avoid possible edge effects around the margin between different tissue types, all voxels with a probability value < 0.2 (absolute threshold; range 0-1) for both GM and WM were excluded. For statistical analysis, a two-sample *t*-test was conducted to identify significant differences in GMV and WMV between MDD patients and HCs, respectively, and the TFCE method with 5000 permutation tests was used for correcting multiple comparisons. The resulted TFCE-statistic map was thresholded at *p* < 0.05, FWE-corrected.

##### Surface-based morphometry analysis.

The synthetic T1-weighted images which generated from mrQ analysis pipeline were then processed for SBM analysis by using Freesurfer v5.3.0 (<http://surfer.nmr.mgh.harvard.edu>) with standard recon-all procedures. The Freesurfer pipeline performs the volumetric GM and WM segmentations, providing several automated cortical parcellations and assigning a neuroanatomical label to each location based on prior atlas. The processing steps included skull stripping, atlas registration, spherical surface registration and parcellation. In this study, cortical parcellation and subcortical segmentation were based on Freesurfer built-in atlas (11, 12), resulting in a total of 82 distinct brain regions, including 74 cortical, seven subcortical regions (the thalamus, caudate, putamen, pallidum, hippocampus, amygdala and nucleus accumbens) per hemisphere and the brainstem. The resulted segmented brain tissues were used for subsequent region of interest (ROI) analysis of qMRI modality. After the recon-all procedures of all participants, cortical thickness maps were smoothed using Gaussian Kernel with FWHM of 10 mm. The vertex-wise group comparison for cortical thickness were performed in Freesurfer using general linear model (GLM) with age, gender and years of education as covariates. All vertex-wise results were corrected for multiple comparisons and separate hemisphere testing using Monte Carlo simulation method with 10000 iterations. The vertex-wise threshold was set at 2, corresponding to *p* < 0.01, and cluster-wise statistical significance was set at corrected *p* < 0.05. A statistical thickness difference map was constructed using −log10 (*p*). Cluster-wise *p* values (CWP) were reported. The volumes of subcortical regions were automatically derived from Freesurfer’s output report for group comparison.

#### Conjunction analysis

A conjunction analysis was conducted to identify regions that showed commonly alternations across all modalities, including structural connectivity, VBM and cortical thickness. Results from different analyses were integrated into AAL atlas based on the MNI coordinates of the peak voxel. The cortical regions were determined based on the consistent alternations in analyses of structural connectivity, VBM and thickness, while the subcortical regions were identified according to analyses of structural connectivity and VBM. Regions especially those play key roles within the LCSPT circuit were included and then defined as ROIs for subsequent qMRI analysis.

#### Quantitative MRI data analysis

##### Data preprocessing

Both the SPGE and SEIR scans were processed by using the mrQ software package (<https://github.com/mezera/mrQ>) in MATLAB to produce the evaluation of MTV and quantitative T1 maps for each participant. The RF coil bias was corrected by combining SPGE scans with a set of low-resolution unbiased SEIR-EPI scans, producing accurate proton density (PD) and unbiased T1 fits across the brain. MTV maps were estimated by calculating the fraction of non-water volume in each voxel while CSF voxels were approximated as entirely filled with water (1). The synthetic T1-weighted images which were spatially matched with MTV maps and optimized for segmentation of brain tissues, were also computed by using mrQ package, and then processed for ROI analysis of qMRI data by using Freesurfer (see **Surface-based morphometry analysis**).

##### ROI analysis for qMRI data.

ROIs were segmented and extracted from Freesurfer’s standard pipeline for each participant and saved as volumetric binary masks. All of these masks were generated in native MTV space and matched well with participants’ synthetic T1w image through manual inspection. The individual ROI mask was then applied on corresponding MTV and T1 maps for each participant, and average MTV and T1 values across voxels within each ROI were computed.

##### Correlation analysis of qMRI measurements.

Regions with group differences in MTV/T1 values were included for this analysis. The GMV and FA values within each region were respectively correlated with its MTV/T1 values across two group. Briefly, each region was defined as a ROI using volumetric 1-mm-diameter sphere centered on its peak MNI coordinates with the strongest statistical significance based on the results of VBM analysis. To extract FA values for each ROI, FA maps of all participants were aligned onto the FMRIB58_FA_1mm template in standard space by using nonlinear registration (13). The ROIs were then applied to normalized FA maps for all participants. The FA values across voxels within each sphere ROI were averaged and extracted. The GMV values within each ROI were averaged and extracted from the preprocessed GM volumes generated from VBM analysis for all participants. The Pearson’s correlation coefficient (*r*) and *p* value between MTV/T1 and the GMV and FA values for each region were calculated. Statistical significance was set at *p* < 0.05, FWE corrected.

#### Supplementary structural network analysis

A supplementary structural network analysis was performed to evaluated the effect of participants’ demographic data on our dMRI results. The procedures performed in the supplementary analysis were similar with the structural network analysis mentioned above (see **Network analysis**) but GLM was established with participants’ age, gender and education years as covariates followed by *t*-test to perform group comparison.

#### Supplementary VBM analysis

Since there were significant differences in education years between two groups, we additionally conducted a supplemental VBM analysis to remove the effect of participants’ demographic data on GMV/WMV measures such that further validate the outcome of our study. The procedures of the supplemental VBM analysis were similar with those mentioned above (see **Voxel-based morphometry analysis**) but GLM rather than two-sample *t*-test was used for statistical analysis. By using GLM, whole-brain GMV/WMV was compared between the patients with MDD and HCs while the demographic data (age, gender and education years) of all participants were classified as covariates and subsequently regressed out.

### Supplementary Results

#### Alternations of structural connectivity in MDD patients

The increased structural connectivity in MDD patients comprised of 215 edges that connected to 76 nodes (Figure 2A), in which the bilateral putamen, thalamus, dorsolateral superior frontal gyrus, superior parietal gyrus, precuneus, right hippocampus (HIP.R) and right caudate (CAU.R) exhibited more than ten increased inter-regional connections. The decreased structural connectivity in MDD patients consisted of 44 edges that linked to 38 nodes (Figure 2B), in which the right thalamus (THA.R), CAU.R, right median cingulate and paracingulate gyri (DCG.R), right anterior cingulate and paracingulate gyri (ACG.R), left insula (INS.L) and fusiform gyrus (FFG.L) were involved in most of decreased connections. At the large-scale networks level, we found that the increased between-network connectivity was expressed largely between the subcortical network and nodes of the default mode network (DMN), while the decreased between-network connectivity was mainly within the subcortical network, DMN, salience/ventral attention network (SN/VAN) and limbic network (LN). Specifically, the increased connectivity within the subcortical regions was observed between the HIP.R and left putamen (PUT.L; *t =* 3.56), the left thalamus (THA.L) and right putamen (PUT.R; *t* = 3.39), the HIP.R and left amygdala (AMYG.L; *t* = 3.32), the PUT.L and right pallidum (PAL.R; *t* = 3.31), the AMYG.L and left caudate (CAU.L; *t* = 3.26), the left hippocampus (HIP.L) and PUT.R (*t* = 3.31) and the HIP.R and THA.L (*t* = 2.91). The increased between-network connectivity was expressed largely between the subcortical regions and nodes of the DMN, specifically the THA.L and left dorsolateral superior frontal gyrus (SFGdor.L; *t* = 3.24), the THA.L and right dorsolateral superior frontal gyrus (SFGdor.R; *t* = 2.67), the THA.R and SFGdor.L (*t* = 2.70), the CAU.R and SFGdor.R (*t* = 3.38) and right medial superior frontal gyrus (SFGmed.R; *t* = 3.31), the PUT.R and SFGdor.L (*t* = 3.88), SFGdor.R (*t* = 3.79) and right inferior orbitofrontal gyrus (ORBinf.R; *t* = 3.45) and between the HIP.R and SFGdor.R (*t* = 2.74). On the other hand, the decreased connectivity within the subcortical regions was detected in the connections of the HIP.R with the PUT.R (*t* = 3.23) and PAL.R (*t* = 2.94), and in the connections of the THA.R with the right parahippocampal gyrus (PHG.R; *t* = 3.64) and CAU.L (*t* = 3.45). The decreased between-network connectivity was mainly between the left superior medial orbitofrontal gyrus (ORBsupmed.L) of the DMN and the left superior orbitofrontal gyrus (ORBsup.L) of the LN (*t* = 4.18), and between the DCG.R of the SN/VAN and the CAU.R (*t* = 3.78) and THA.R (*t* = 2.88) of the subcortical network. Notably, increased inter-regional connections accounted for the majority among those hub regions, particularly in which the PUT.L, right superior parietal gyrus (SPG.R) and THA.L only exhibited increased connections.

#### Alternations of the thalamus-centered structural connectivity in MDD patients

To identify the specific pattern of thalamic structural connectivity in MDD patients, we extracted the bilateral thalamus-to-whole-brain connections identified by the NBS analysis (supplementary Figure S1A). These connections formed a thalamus-centered network, primarily involving the bilateral supplementary motor area, dorsolateral superior frontal gyrus, putamen, precuneus, superior parietal gyrus, superior occipital gyrus and middle frontal gyrus. Specifically, THA.L-centered network comprised of 15 increased connections with 15 nodes, mainly involving the left supplementary motor area (SMA.L), left paracentral lobule (PCL.L), PUT.R, SFGdor.L, SPG.R and left precuneus (PCUN.L). The THA.R-centered network comprised of 20 edges connecting to 20 nodes, including 16 increased connections and four decreased connections. The increased connections linked the THA.R with the bilateral supplementary motor area, right superior occipital gyrus (SOG.R), right postcentral gyrus (PoCG.R), right precentral gyrus (PreCG.R) and PUT.L whereas the decreased connections linked the THA.R with the PHG.R, CAU.L, DCG.R and PAL.R. These connections remained significant at higher supra-threshold of *t* > 3.0. The results of the thalamus-whole brain fiber tracking showed a significant hyper-connectivity pattern in MDD patients compared with HCs after TFCE correction (supplementary Figure S1B). We found that the target regions structurally connecting to the thalamus were located in the cortical regions e.g., the bilateral middle frontal gyrus, middle temporal gyrus, dorsolateral superior frontal gyrus, precentral and postcentral gyrus, precuneus, inferior temporal gyrus, superior temporal gyrus, superior medial frontal gyrus, insula, inferior orbitofrontal gyrus and anterior cingulate and paracingulate gyri as well as subcortical regions including the bilateral putamen, hippocampus, parahippocampal gyrus, caudate and pallidum. No significant decreased structural connectivity was survived after multiple comparison correction in MDD patients relative to HCs.

#### Results of conjunction analysis

A total of five regions including one cortical and four subcortical regions were identified (supplementary Table S5). These regions have shown to play key roles within the LCSPT circuit but involved in cross-modal structural alternations (e.g., structural connectivity and morphologic features) in patients with MDD.

#### The correlation of qMRI measurements across 2 groups

The correlation analysis showed that T1 values were negatively correlated with corresponding GMV values in the THA.L (*r* = -0.349, *p* = 0.002; supplementary Figure S2) across two groups.

#### Results of supplementary structural network analysis

By using NBS analysis, we only identified a single network after correcting for all the covariates (supplementary Figure S3). The identified network consisted of 212 edges connecting to 75 nodes, representing increased structural connectivity in MDD patients (supplementary Figure S3A). Among these nodes, the bilateral putamen, thalamus, dorsolateral superior frontal gyrus, hippocampus, superior parietal gyrus, right inferior temporal gyrus (ITG.R), CAU.R, SMA.L, SOG.R, PCUN.L and right middle temporal gyrus (MTG.R) involved ten and more increased inter-regional connections, which were consistent with the results in the main text. Similarly, the identified increased connectivity was further classified into seven-network parcellation with 100 parcels. We found the most of increased connections were still within the subcortical regions and between the subcortical network and other cortical network. Specifically, the increased connections within the subcortical regions were observed between the PUT.R and HIP.L (*t* = 3.37), the PUT.R and THA.L (*t* = 3.26), the PUT.L and HIP.R (*t* = 3.26), the PUT.L and PUT.R (*t* = 3.10), the PUT.L and PAL.R (*t* = 3.08), the PUT.L and THA.R (*t* = 2.94), the HIP.R and AMYG.L (*t* = 2.93), the AMYG.L and CAU.L (*t* = 2.78) and the HIP.R and THA.L (*t* = 2.55). The between-network connections were mainly detected between the subcortical regions and regions within the somatomotor network, specifically between the PUT.L and left precentral gyrus (PreCG.L; *t* = 3.50), the PUT.L and PCL.L (*t* = 3.49), the THA.L and PCL.L (*t* = 3.23), the PUT.R and PoCG.R (*t* = 3.08), the PUT.L and left postcentral gyrus (PoCG.L; *t* = 3.08), the CAU.L and right superior temporal gyrus (STG.R; *t* = 3.02) and the PUT.R and PreCG.R (*t* = 3.00). These supplementary results revealed a similar increased connectivity pattern with those in the main text in MDD patients, both indicating the subcortical regions may be closely associated with disrupted structural connectivity in MDD patients. The supplementary results suggest altered structural connectivity in MDD patients are unlikely to be effect by participants’ age, gender and education years.

#### Results of supplementary VBM analysis

The results of supplementary VBM analysis were comparable to those in the main text (supplementary Figure S4 and Table S6). A total of 23 clusters were identified showing significant reduced GMV in patients with MDD relative to HCs, while WMV reductions were found in eight clusters (supplementary Table S6). Additionally, reduced GMV values of the key regions e.g., the bilateral thalamus, bilateral insula, left parahippocampal gyrus (PHG.L), right amygdala (AMYG.R), left inferior orbitofrontal gyrus (ORBinf.L) and ORBsup.L were still observed even participants’ demographic data were regressed out. The GMV values of these key regions were also extracted from the identified clusters for visualization (supplementary Figure S4B). The supplementary results of VBM analysis indicate that the overall volume reduction in MDD patients is unlikely to be induced by participants’ demographic data. Notably, there was still no significant cluster showing larger GMV/WMV in MDD patients compared with HCs.

**Table S1.** Demographic and clinical characteristics of participants.

|  | **MDD (n=56)** | | **HC (n=35)** | |
| --- | --- | --- | --- | --- |
|  | Mean | SD | Mean | SD |
| Years of age | 36.36 | 15.96 | 39.40 | 14.77 |
| Years of education | 12.82 | 3.17 | 14.94 | 2.84 |
| HAMD | 19.63 | 7.61 |  |  |
| HAMA | 16.23 | 6.77 |  |  |
|  | N | % | N | % |
| Male | 15 | 27 | 13 | 37 |
| Multiple depressive episodes | 22 | 39 |  |  |
| Psychiatric comorbidities | 7 | 13 |  |  |
| Current medications | 48 | 86 |  |  |
| Duration exposed to medications |  |  |  |  |
| < 3 months | 24 | 43 |  |  |
| ≥ 3 months | 24 | 43 |  |  |

Note: MDD, major depression disorder; HC, healthy control; HAMD, Hamilton Depression Rating Scale; HAMA, Hamilton Anxiety Scale.

**Table S2.** Abbreviations of regions from AAL atlas.

| **Index** | **Regions** | **Abbreviation** |
| --- | --- | --- |
| 1 | Precentral gyrus | PreCG.L |
| 2 | Precentral gyrus | PreCG.R |
| 3 | Superior frontal gyrus, dorsolateral | SFGdor.L |
| 4 | Superior frontal gyrus, dorsolateral | SFGdor.R |
| 5 | Superior frontal gyrus, orbital part | ORBsup.L |
| 6 | Superior frontal gyrus, orbital part | ORBsup.R |
| 7 | Middle frontal gyrus | MFG.L |
| 8 | Middle frontal gyrus | MFG.R |
| 9 | Middle frontal gyrus, orbital part | ORBmid.L |
| 10 | Middle frontal gyrus, orbital part | ORBmid.R |
| 11 | Inferior frontal gyrus, opercular part | IFGoperc.L |
| 12 | Inferior frontal gyrus, opercular part | IFGoperc.R |
| 13 | Inferior frontal gyrus, triangular part | IFGtriang.L |
| 14 | Inferior frontal gyrus, triangular part | IFGtriang.R |
| 15 | Inferior frontal gyrus, orbital part | ORBinf.L |
| 16 | Inferior frontal gyrus, orbital part | ORBinf.R |
| 17 | Rolandic operculum | ROL.L |
| 18 | Rolandic operculum | ROL.R |
| 19 | Supplementary motor area | SMA.L |
| 20 | Supplementary motor area | SMA.R |
| 21 | Olfactory cortex | OLF.L |
| 22 | Olfactory cortex | OLF.R |
| 23 | Superior frontal gyrus, medial | SFGmed.L |
| 24 | Superior frontal gyrus, medial | SFGmed.R |
| 25 | Superior frontal gyrus, medial orbital | ORBsupmed.L |
| 26 | Superior frontal gyrus, medial orbital | ORBsupmed.R |
| 27 | Gyrus rectus | REC.L |
| 28 | Gyrus rectus | REC.R |
| 29 | Insula | INS.L |
| 30 | Insula | INS.R |
| 31 | Anterior cingulate and paracingulate gyri | ACG.L |
| 32 | Anterior cingulate and paracingulate gyri | ACG.R |
| 33 | Median cingulate and paracingulate gyri | DCG.L |
| 34 | Median cingulate and paracingulate gyri | DCG.R |
| 35 | Posterior cingulate gyrus | PCG.L |
| 36 | Posterior cingulate gyrus | PCG.R |
| 37 | Hippocampus | HIP.L |
| 38 | Hippocampus | HIP.R |
| 39 | Parahippocampal gyrus | PHG.L |
| 40 | Parahippocampal gyrus | PHG.R |
| 41 | Amygdala | AMYG.L |
| 42 | Amygdala | AMYG.R |
| 43 | Calcarine fissure and surrounding cortex | CAL.L |
| 44 | Calcarine fissure and surrounding cortex | CAL.R |
| 45 | Cuneus | CUN.L |
| 46 | Cuneus | CUN.R |
| 47 | Lingual gyrus | LING.L |
| 48 | Lingual gyrus | LING.R |
| 49 | Superior occipital gyrus | SOG.L |
| 50 | Superior occipital gyrus | SOG.R |
| 51 | Middle occipital gyrus | MOG.L |
| 52 | Middle occipital gyrus | MOG.R |
| 53 | Inferior occipital gyrus | IOG.L |
| 54 | Inferior occipital gyrus | IOG.R |
| 55 | Fusiform gyrus | FFG.L |
| 56 | Fusiform gyrus | FFG.R |
| 57 | Postcentral gyrus | PoCG.L |
| 58 | Postcentral gyrus | PoCG.R |
| 59 | Superior parietal gyrus | SPG.L |
| 60 | Superior parietal gyrus | SPG.R |
| 61 | Inferior parietal, but supramarginal and angular gyri | IPL.L |
| 62 | Inferior parietal, but supramarginal and angular gyri | IPL.R |
| 63 | Supramarginal gyrus | SMG.L |
| 64 | Supramarginal gyrus | SMG.R |
| 65 | Angular gyrus | ANG.L |
| 66 | Angular gyrus | ANG.R |
| 67 | Precuneus | PCUN.L |
| 68 | Precuneus | PCUN.R |
| 69 | Paracentral lobule | PCL.L |
| 70 | Paracentral lobule | PCL.R |
| 71 | Caudate nucleus | CAU.L |
| 72 | Caudate nucleus | CAU.R |
| 73 | Lenticular nucleus, putamen | PUT.L |
| 74 | Lenticular nucleus, putamen | PUT.R |
| 75 | Lenticular nucleus, pallidum | PAL.L |
| 76 | Lenticular nucleus, pallidum | PAL.R |
| 77 | Thalamus | THA.L |
| 78 | Thalamus | THA.R |
| 79 | Heschl gyrus | HES.L |
| 80 | Heschl gyrus | HES.R |
| 81 | Superior temporal gyrus | STG.L |
| 82 | Superior temporal gyrus | STG.R |
| 83 | Temporal pole: superior temporal gyrus | TPOsup.L |
| 84 | Temporal pole: superior temporal gyrus | TPOsup.R |
| 85 | Middle temporal gyrus | MTG.L |
| 86 | Middle temporal gyrus | MTG.R |
| 87 | Temporal pole: middle temporal gyrus | TPOmid.L |
| 88 | Temporal pole: middle temporal gyrus | TPOmid.R |
| 89 | Inferior temporal gyrus | ITG.L |
| 90 | Inferior temporal gyrus | ITG.R |

The brain regions were defined by Tzourio-Mazoyer et al. (2002)

**Table S3.** Clusters showed significant GMV/WMV differences between MDD patients and HCs.

| **Anatomical region ^a^** | **Side** | **MNI coordinates ^b^** | | | **Cluster size** | ***p* value**  **(FWE-corrected)** |
| --- | --- | --- | --- | --- | --- | --- |
|  |  | x | y | z |  |  |
| **GMV** | | | | | | |
| THA | L | -8 | -6 | 4 | 1112 | <0.001 |
| AMYG | R | 21 | 0 | -15 | 353 | 0.001 |
| HIP | R | 18 | -9 | -15 | 265 | 0.001 |
| PHG | L | -20 | -26 | -18 | 516 | 0.002 |
| PHG | R | 14 | 0 | -18 | 407 | 0.002 |
| THA | R | 10 | -9 | 7 | 328 | 0.005 |
| STG | R | 65 | -20 | 12 | 2480 | 0.005 |
| INS | L | -32 | 24 | 0 | 1345 | 0.005 |
| HES | R | 39 | -24 | 9 | 528 | 0.005 |
| ORBinf | L | -21 | 18 | -14 | 583 | 0.006 |
| PUT | L | -24 | 17 | 6 | 187 | 0.006 |
| REC | R | 2 | 51 | -18 | 551 | 0.007 |
| OLF | L | -20 | 6 | -12 | 391 | 0.007 |
| SMG | R | 56 | -33 | 27 | 672 | 0.008 |
| INS | R | 39 | 11 | 2 | 2259 | 0.009 |
| REC | L | 0 | 49 | -18 | 925 | 0.009 |
| ORBsupmed | R | 9 | 51 | -12 | 768 | 0.009 |
| ACG | L | -4 | 35 | -9 | 423 | 0.009 |
| MTG | R | 66 | -22 | -3 | 160 | 0.010 |
| STG | L | -60 | -17 | 9 | 1731 | 0.017 |
| MFG | L | -33 | 50 | 1 | 280 | 0.017 |
| HES | L | -51 | -16 | 10 | 299 | 0.018 |
| TPOmid | R | 21 | 12 | -36 | 522 | 0.019 |
| ORBmid | L | -39 | 45 | 0 | 193 | 0.019 |
| FFG | R | 28 | 5 | -41 | 687 | 0.021 |
| SFGdor | L | -16 | 50 | 34 | 808 | 0.027 |
| MTG | L | -63 | -33 | 2 | 382 | 0.035 |
| **WMV** | | | | | | |
| THA | R | 10 | -22 | 9 | 613 | 0.004 |
| THA | L | -6 | -15 | 10 | 431 | 0.006 |
| PreCG | R | 32 | -19 | 57 | 1192 | 0.010 |
| PreCG | L | -28 | -22 | 55 | 625 | 0.015 |
| PoCG | L | -26 | -40 | 48 | 608 | 0.017 |
| SMA | R | 9 | -27 | 58 | 446 | 0.017 |
| PCL | R | 6 | -30 | 61 | 330 | 0.018 |
| PCUN | L | -15 | -42 | 60 | 244 | 0.018 |
| DCG | L | -14 | -16 | 45 | 214 | 0.021 |
| CAU | R | 16 | -18 | 21 | 397 | 0.022 |
| IFGoperc | R | 36 | 8 | 30 | 128 | 0.029 |
| SFGdor | R | 15 | 11 | 54 | 302 | 0.030 |
| PoCG | R | 30 | -27 | 60 | 217 | 0.031 |
| ACG | L | -3 | 35 | -2 | 180 | 0.031 |
| STG | R | 56 | -24 | 1 | 862 | 0.034 |

Note: ^a^ For full names of these abbreviations from AAL atlas, see supplementary Table S2. ^b^ MNI coordinates of the voxel with maximum statistical significance for identified clusters. Each cluster was represented by a coordinate of the peak voxel in this cluster. GMV, gray matter volume; WMV, white matter volume; MDD, major depression disorder; HC, healthy control; L, left; R, right.

**Table S4.** Surface-based cluster summary of significant differences in cortex thickness between MDD patients and HCs.

| **Anatomical location ^a^** | **Side** | **Size (mm^2^)** | **MNI coordinates ^*^** | | | **CWP** |
| --- | --- | --- | --- | --- | --- | --- |
|  |  |  | x | y | z |  |
| MFG | R | 3790.43 | 37.1 | 27.3 | 39.9 | 0.0002 |
| MTG | L | 2277.56 | -62.2 | -21.5 | 1.8 | 0.0002 |
| ORBinf | R | 2141.24 | 43.5 | 33.5 | -13.3 | 0.0002 |
| SFGdor | L | 1739.25 | -19.5 | 10.3 | 56.0 | 0.0002 |
| CAL | L | 1696.76 | -15.4 | -77.3 | 10.9 | 0.0002 |
| SPG | L | 1170.05 | -25.5 | -54.8 | 63.6 | 0.0002 |
| INS | L | 1163.41 | -31.9 | 26.5 | 9.0 | 0.0002 |
| ORBmid | L | 1138.21 | -38.4 | 44.7 | -3.8 | 0.0002 |
| SFGmed | L | 667.90 | -11.7 | 63.0 | 2.5 | 0.0024 |
| ITG | L | 529.68 | -49.8 | -62.8 | -5.9 | 0.014 |
| ITG | L | 502.62 | -50.3 | -48.8 | -17.2 | 0.019 |
| MFG | L | 488.18 | -25.7 | 28.4 | 33.5 | 0.023 |
| SMA | R | 481.17 | 7.4 | 4.5 | 59.7 | 0.030 |
| MTG | R | 471.28 | 60.0 | -15.6 | -16.8 | 0.034 |

Note: ^a^ For full names of these abbreviations from AAL atlas, see supplementary Table S2. ^*^ Coordinates of the peak vertex for the cluster. CWP, cluster-wise *p* value; L, left; R, right.

**Table S5.** Regions with consistent alternations across modalities within the LCSPT circuit in MDD patients.

| **Regions** | **Structural connectivity ^a^** | | **Morphometry measurements ^b^** | | | |
| --- | --- | --- | --- | --- | --- | --- |
|  | Contrast | N | VBM | *p* value | SBM | *p* value |
| **Cortical regions** | | | | | | |
| ORBmid.L | MDD > HC | 2 | GMV | 0.017 | Cortical thickness | 0.0002 |
|  | MDD < HC | 1 |  |  |  |  |
| **Subcortical regions** | | | | | | |
| PUT.L | MDD > HC | 27 | GMV | 0.006 | - | |
| THA.R | MDD > HC | 16 | GMV | 0.005 | - | |
|  | MDD < HC | 4 | WMV | 0.004 | - | |
| THA.L | MDD > HC | 15 | GMV | <0.001 | - | |
|  |  |  | WMV | 0.006 | - | |
| AMYG.R | MDD > HC | 1 | GMV | 0.001 | - | |

Note: ^a^ Structural connectivity corresponds to the results of NBS analysis in dMRI data analysis. ^b^ Significant group differences were only detected in the contrast of MDD < HC in the results of VBM and SBM analysis. N are number of connections. VBM, Voxel-based morphometry; SBM, Surface-based morphometry; MDD, major depression disorder; HC, healthy controls. For full names of these abbreviations from AAL atlas, see supplementary Table S2.

**Table S6.** Clusters showed significant GMV/WMV differences between MDD patients and HCs after controlling for the demographic data.

| **Anatomical region ^a^** | **Side** | **MNI coordinates ^b^** | | | **Cluster size** | ***p* value** |
| --- | --- | --- | --- | --- | --- | --- |
|  |  | x | y | z |  |  |
| **GMV** | | | | | | |
| THA | L | -8 | -10 | 6 | 785 | <0.001 |
| PHG | L | -14 | -34 | -9 | 410 | 0.001 |
| AMYG | R | 22 | 2 | -15 | 313 | 0.002 |
| THA | R | 12 | -9 | 9 | 241 | 0.002 |
| OLF | L | 0 | 8 | -8 | 400 | 0.003 |
| STG | R | 63 | -16 | 10 | 2075 | 0.005 |
| INS | R | 40 | 10 | 0 | 1912 | 0.005 |
| INS | L | -32 | 24 | -2 | 1388 | 0.005 |
| ORBinf | L | -24 | 20 | -15 | 1228 | 0.006 |
| STG | L | -51 | -3 | 0 | 1088 | 0.006 |
| REC | R | 3 | 39 | -23 | 726 | 0.006 |
| TPOsup | L | -22 | 5 | -20 | 356 | 0.006 |
| TPOsup | R | 57 | 3 | 1 | 265 | 0.006 |
| ORBsup | L | -24 | 17 | -12 | 190 | 0.006 |
| IFGoperc | L | -56 | 11 | 6 | 485 | 0.007 |
| REC | L | 0 | 56 | -20 | 1057 | 0.008 |
| IFGtriang | L | -51 | 17 | 0 | 775 | 0.009 |
| PoCG | L | -26 | -46 | 55 | 325 | 0.019 |
| IPL | L | -33 | -51 | 54 | 1133 | 0.021 |
| FFG | R | 42 | -16 | -30 | 276 | 0.027 |
| SMG | L | -62 | -34 | 39 | 119 | 0.030 |
| MFG | L | -24 | 41 | 28 | 300 | 0.032 |
| SFGdor | L | -16 | 50 | 34 | 229 | 0.033 |
| **WMV** | | | | | | |
| PreCG | R | 18 | -24 | 67 | 1178 | 0.004 |
| THA | L | -6 | -15 | 10 | 360 | 0.004 |
| THA | R | 8 | -18 | 12 | 484 | 0.006 |
| PoCG | L | -26 | -40 | 49 | 582 | 0.008 |
| STG | R | 44 | -31 | 3 | 414 | 0.009 |
| PreCG | L | -28 | -22 | 55 | 577 | 0.011 |
| PCL | R | 6 | -31 | 61 | 310 | 0.013 |
| DCG | L | -14 | -15 | 46 | 225 | 0.020 |

Note: ^a^ For full names of these abbreviations from AAL atlas, see supplementary Table S2. ^b^ MNI coordinates of the voxel with maximum statistical significance for identified clusters. Each cluster was represented by a coordinate of the peak voxel in this cluster. GMV, gray matter volume; WMV, white matter volume; MDD, major depression disorder; HC, healthy control; L, left; R, right.

**Figure S1. Significant altered structural connectivity and connectivity patterns in the thalamus-centered network.**

(A) Bilateral thalamus-centered networks identified by the whole-brain NBS analysis were additionally extracted and visualized. The nodes with increased and decreased connections are shown in orange and blue, respectively. Size of nodes indicate *t* values of edge between the thalamus and target nodes. The bilateral thalamus nodes are emphasized in red. The edges are shown in the same color as their connected nodes with the same widths. Nodes with most of the inter-regional connections are labelled. (B) Significant group differences in the bilateral thalamus-whole brain tractography. The color bar indicates 1-*p* value with a significant threshold of 0.95, corresponding to *p* < 0.05, FWE-corrected. Representative coronal, sagittal and axial slices of the significant connections were overlaid on a standard fractional anisotropy template from FSL package. For abbreviation and index of AAL regions, see supplementary Table S2.

**Figure S2. Correlation between T1 and GMV values in the THA.L across 2 groups.**

**Figure S3. Significant altered structural connectivity in MDD patients with all covariates removed and an increased connectivity pattern among 7 networks and subcortical networks.**

(A) Increased structural connectivity in MDD patients compared with HCs. (B) The circle plot depicting the increased connectivity pattern within and between 7 network and subcortical networks. For abbreviation and index of AAL regions, see supplementary Table S2.

**Figure S4. The results of supplementary VBM analysis with covariates removed and between-group differences of the GMV values in specific regions.**

(A) GMV (upper) and WMV (lower) reductions in MDD patients compared with HCs. The color bar corresponds to FWE-corrected *p* < 0.05 and lower. (B) Violin plots depicting group differences of the GMV values within 8 key regions. Since the raw statistic map was transformed into TFCE values after TFCE method, the GMV values of these regions were extracted from the group-comparison statistic map with threshold of *p* < 0.001 (uncorrected) at the voxel level. For abbreviation of AAL regions, see supplementary Table S2.
